# Controlling the Transmission Dynamics of HAT Incorporating Impacts of Temperature

**DOI:** 10.64898/2026.01.29.26345117

**Authors:** Anthony Okumu, Nicholas Kwasi-Do Ohene Opoku

## Abstract

Human African Trypanosomiasis (HAT) remains a persistent public health threat in sub-Saharan Africa, with transmission dynamics tightly coupled to the ecology and physiology of its tsetse fly vector. Despite growing evidence that temperature strongly modulates vector survival, development, and biting behavior, most existing transmission models assume static environmental conditions. We develop a model for HAT that incorporates temperature-dependent vector recruitment, mortality, and biting rates, thereby mechanistically linking environmental variability to epidemiological outcomes. The model couples human and tsetse populations and admits both disease-free and endemic equilibria. Using the next-generation matrix approach, we derive an explicit expression for the basic reproduction number and show that it depends nonlinearly on temperature through multiple entomological pathways. Bifurcation analysis reveals a forward transcritical bifurcation, indicating a clear threshold for disease persistence. Our findings demonstrate how temperature can fundamentally alter transmission potential and control thresholds, highlighting the importance of integrating climate-sensitive vector biology into HAT risk assessment and intervention planning under ongoing environmental change.

## 1 Introduction

Human African Trypanosomiasis (HAT), or sleeping sickness, is a vector-borne parasitic disease caused by Trypanosoma brucei subspecies [1]. Two forms of the disease are recognized; the acute form caused by T. b. rhodesiense, prevalent in Eastern and Southern Africa, and the chronic form caused by T. b. gambiense, accounting for over 85% of cases in Western and Central Africa [2]. Transmission occurs via the bite of infected tsetse flies (Glossina spp.), which acquire the parasite from human or animal reservoirs [2]. Once infected, the fly becomes a lifelong vector, with the parasite undergoing development in the midgut before migrating to the salivary glands to become infective [3]. In humans, infection progresses from systemic symptoms to neurological involvement, often leading to coma and death if untreated [4]. The spatial distribution of HAT is shaped by the ecology of tsetse flies and the presence of reservoirs. T. b. gambiense is primarily anthroponotic, while T. b. rhodesiense is zoonotic, maintained in wildlife and livestock [5]. Environmental factors, particularly vegetation, humidity, and temperature, play a critical role in the survival and reproduction of tsetse flies. These flies reproduce slowly through a unique process called adenotrophic viviparity, in which the female nourishes a single developing larva internally and deposits it every 9 − 10 days [6]. After pupation in the soil, the timing and success of adult emergence are highly sensitive to temperature. High temperatures accelerate development but can also be lethal; sustained exposure to higher temperatures significantly reduces pupal survival and prevent emergence entirely [7]. The epidemiology of HAT is linked to the ecological and physiological characteristics of its vector, Glossina spp., whose survival, development, and vectorial capacity are highly sensitive to environmental conditions, particularly temperature. As global climate variability intensifies, understanding and quantifying the temperature-mediated dynamics of tsetse populations and their implications for HAT transmission has become increasingly critical. Understanding how climatic factors, particularly temperature, affect HAT population dynamics is essential for effective control of HAT transmission, especially in the context of environmental change.

One of the foundational efforts in this domain was presented by [8], who developed a temperature-dependent model to estimate the climatic suitability for HAT transmission. Their framework integrated thermal thresholds affecting tsetse biology and host–vector interactions, identifying a temperature window of 20.7^°^*C* to 26.1^°^*C* as optimal for transmission. While the model successfully demonstrated the sensitivity of transmission potential to temperature, it did not consider broader climatic drivers, control interventions, or the epidemiological consequences of temperature-induced shifts in vector dynamics, particularly the basic reproduction number, ℛ_0_.

Subsequent work by [9] introduced an agent-based model incorporating seasonal variability and an explicit representation of the tsetse life cycle as an epidemiological class. Their simulations effectively captured the spatiotemporal dynamics of T. b. rhodesiense transmission and provided insights into risk stratification among human subpopulations. However, the model did not assess how temperature affects vector development or mortality, nor did it incorporate analytical tools such as optimal control or sensitivity analysis to guide targeted intervention strategies. [10] contributed a biologically detailed model focusing on temperature-dependent larval development and pupal mortality in Glossina. Using empirical data, they parameterized vector life stages under variable temperature regimes. Though this work advanced the entomological modeling of tsetse ecology, it did not extend to the human epidemiological domain, thereby missing the crucial interface between vector dynamics and HAT transmission.

[6] employed a species distribution modeling approach to predict shifts in Glossina habitat suitability under projected climate scenarios. Their work provided empirical evidence that rising temperatures are likely to reshape tsetse distribution, with significant implications for the geographic footprint of HAT. However, the model lacked an epidemiological structure and did not link vector distribution to transmission potential or assess intervention efficacy under changing environmental conditions. Mathematical models incorporating disease control strategies have also been developed. [11] proposed a compartmental model including chemotherapeutic and vector control interventions, with stability and threshold analysis. While this framework allowed for the computation of ℛ_0_ and optimal control strategies, the model assumed static environmental conditions and did not account for temperature-sensitive vector biology. Similarly, [12] applied Pontryagin’s Maximum Principle to evaluate combined control measures, showing that treatment, personal protection, and insecticide application synergistically reduce transmission. However, their model treated all biological parameters as temperature-invariant, thus omitting the substantial influence of thermal variability on vector survival, biting rate, and reproductive potential. [13] introduced relapse dynamics and health systems factors into a deterministic HAT model, identifying healthcare access, health education, and vaccination as key levers in reducing transmission. Although comprehensive in addressing human behavioral and systemic determinants, the model remained environmentally agnostic. In parallel, [14] presented a mechanistic model examining the influence of rising temperatures on Glossina pallidipes populations in Zimbabwe. Their findings suggested altitudinal range shifts in tsetse distribution but lacked an epidemiological coupling to human disease outcomes or intervention scenarios. Collectively, these studies underscore the biological and epidemiological importance of temperature in shaping HAT transmission. However, a common limitation across the existing literature is the incomplete integration of climate-sensitive entomological parameters within a formal disease transmission framework. Specifically, models rarely incorporate temperature-dependent functions for key vector parameters, such as biting rate, transmission, and mortality, and even fewer assess the implications of such variability on transmission thresholds, intervention efficacy, or long-term disease persistence. To address these deficiencies, the present study develops a deterministic HAT transmission model that explicitly incorporates temperature-dependent vector parameters. By embedding thermal sensitivity into both vector demography and vector–host transmission dynamics, the model provides a mechanistic link between environmental variability and epidemiological outcomes.

## 2 Model Formulation

Transmission of HAT results from interactions between human hosts and tsetse fly vectors, with disease persistence governed by the coupled dynamics of both populations[15, 16]. To describe this process, the total human population *N*_H_(*t*) is divided into susceptible *S*_H_(*t*), exposed *E*_H_(*t*), infectious *I*_SH_(*t*), and recovered *R*_H_(*t*) classes, based on the natural history of HAT, which includes a latent period prior to infectiousness[11]. The total tsetse fly population *N*_v_(*t*) is divided into susceptible *S*_v_(*t*), exposed *E*_v_(*t*), and infectious *I*_v_(*t*) compartments, reflecting the extrinsic incubation period of *Trypanosoma* parasites in the vector[15]. HAT transmission occurs through a cyclical host-vector process in which tsetse flies act as obligate biological vectors of *Trypanosoma* spp. parasites [17]. Direct human-to-human transmission is assumed negligible, consistent with epidemiological evidence [17]. Thus, susceptible humans enter the population at a constant rate *λ*_H_ and infection of humans occurs exclusively through the bite of an infectious tsetse fly, as no alternative transmission routes play a significant epidemiological role [17]. The biting rate of tsetse flies, denoted *b*(*T*), is assumed to depend on ambient temperature *T*, reflecting experimental and field evidence that temperature influences vector activity, feeding frequency, and host-vector contact rates [18]. Given a bite from an infectious vector, the probability of successful transmission to a susceptible human is denoted by *p*_H_. The force of infection acting on the human population is therefore defined as

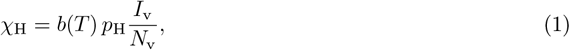

representing the per capita rate at which susceptible humans acquire infection following contact with infectious vectors. Exposed humans progress to the infectious class at rate *γ*, reflecting the incubation period of HAT as explained by [11]. Infectious individuals recover following effective treatment at rate *δ* or experience disease-induced mortality at per capita rate *θ* in the absence of treatment [11]. Recovered individuals are assumed to lose protective immunity and return to the susceptible class at rate *τ*, consistent with evidence that infection does not confer long-lasting immunity against reinfection [11]. Natural mortality occurs in all human compartments at per capita rate *µ*_H_.

Tsetse fly recruitment is driven by the maturation of immature stages into adult flies at a temperature-dependent rate *β*_v_(*T*), reflecting the strong influence of temperature on tsetse development and reproduction [19]. Population growth is limited by environmental resources and is therefore assumed to follow logistic dynamics with carrying capacity *K*, a common assumption in vector population modelling [20]. A proportion *η*_v_ of immature flies die before reaching adulthood, while the remaining fraction 1 − *η*_v_ successfully mature, consistent with observed high mortality rates during early life stages of tsetse flies[20]. The recruitment rate of susceptible adult tsetse flies is thus given by

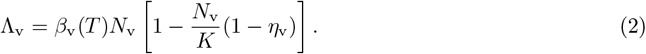

Susceptible tsetse flies acquire infection when feeding on infectious humans. The probability of pathogen transmission from humans to vectors is denoted by *p*_v_, in line with empirical and theoretical studies of HAT transmission efficiency reported by [15]. The force of infection acting on the vector population is therefore

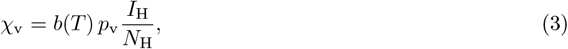

representing the rate at which susceptible vectors become exposed during blood meals. Exposed tsetse flies progress to the infectious class at rate *π*, corresponding to the extrinsic incubation period of *Trypanosoma* parasites, while Infected tsetse flies are assumed not to recover and remain infectious for life, consistent with experimental evidence by [15]. Furthermore, HAT infection is assumed not to induce additional mortality in tsetse flies, as vector survival is primarily governed by environmental conditions rather than pathogen burden[21, 22]. Adult tsetse flies in all epidemiological classes experience natural mortality at a temperature-dependent rate *µ*_v_(*T*), reflecting the well-documented sensitivity of tsetse survival to thermal conditions [19]. Figure 1 depicts the HAT transmission within the human and tsetse fly populations.

**Fig. 1.**
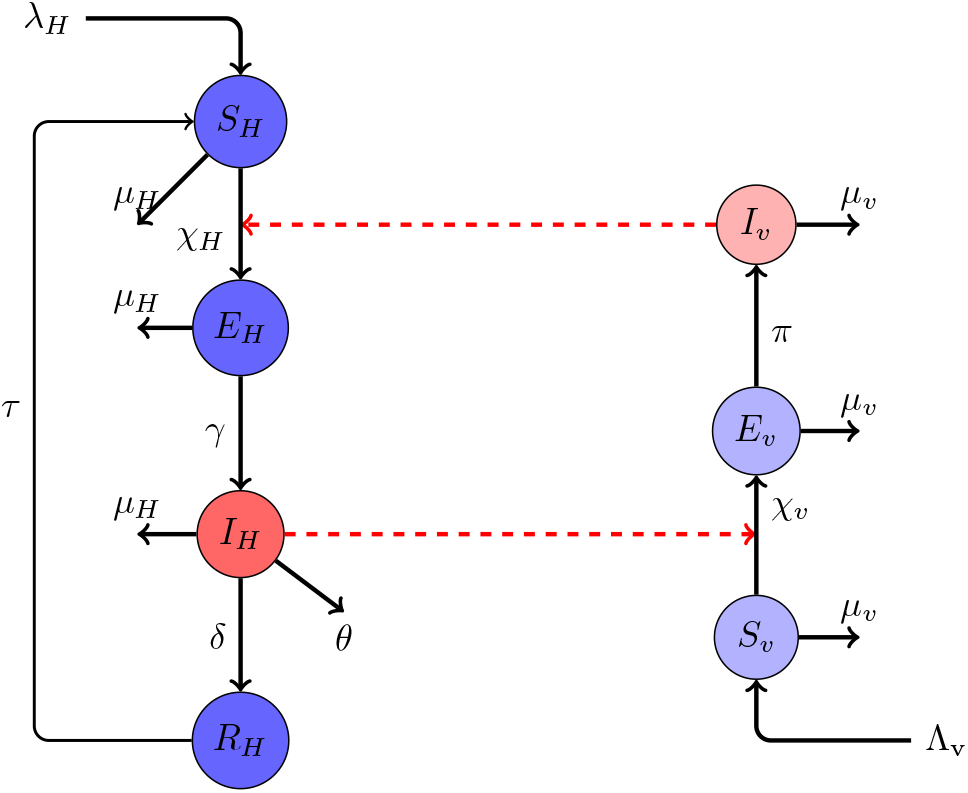
Compartmental diagram of HAT transmission between human and tsetse populations. Red dotted arrow lines indicate the HAT transition path between human and tsetse populations.

The state variables and parameters used in the model are described in Table 1.

**Table 1.**
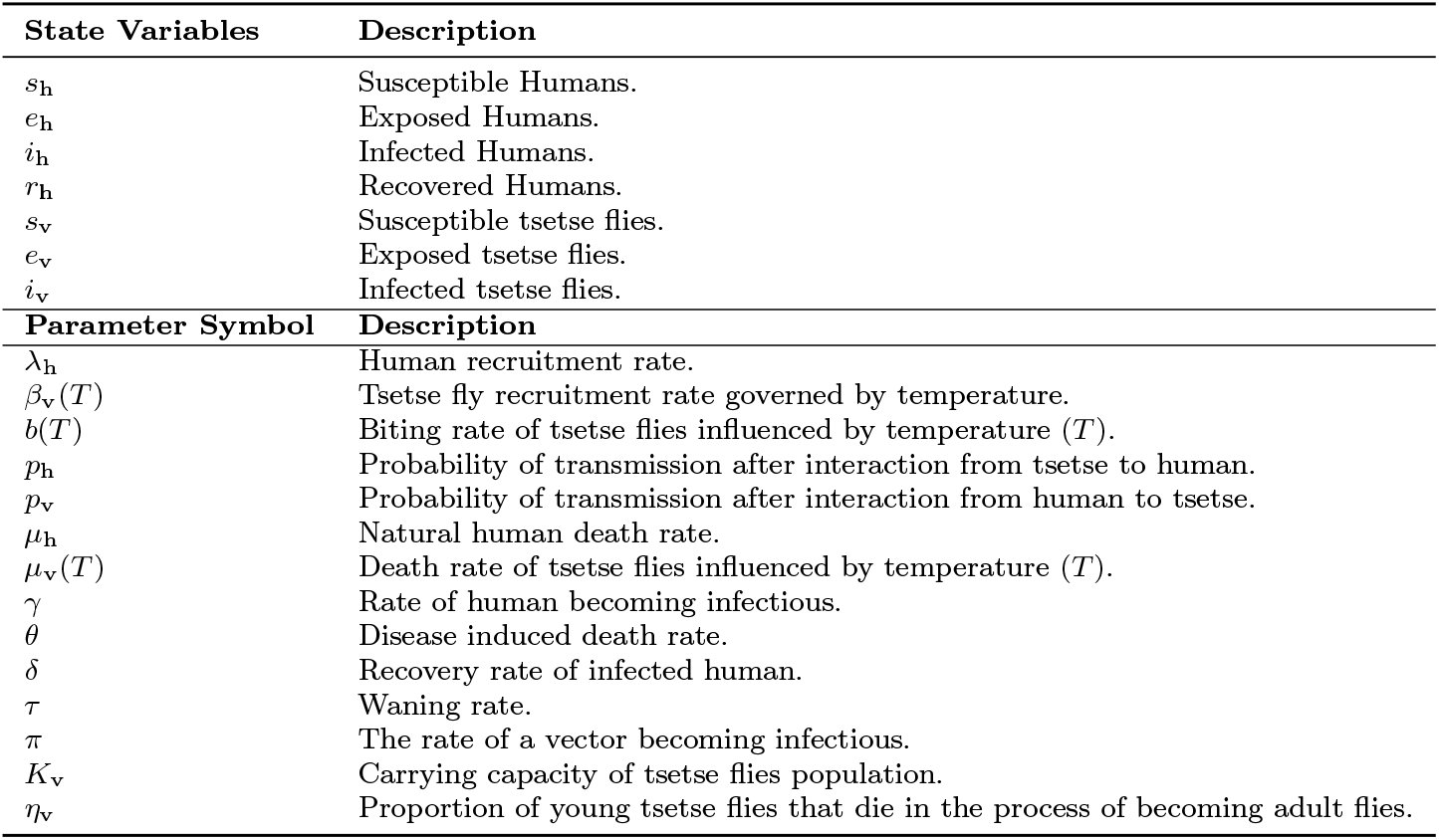
Description of model’s state and parameter variables.

## 3 Model Analysis

System (4) describes the epidemiological interaction between the human host and tsetse vector populations.

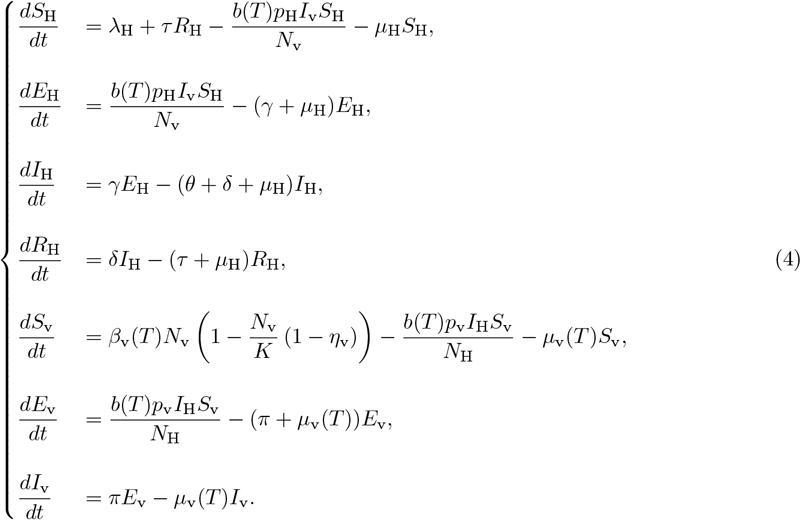

We reformulate System (4) into a more convenient form for further analysis. Let

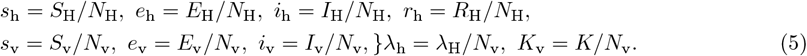

Therefore,

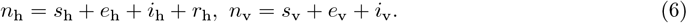

Using the transformations in Equations (5) and (6), System (4) is rewritten into an equivalent dimensionless system as

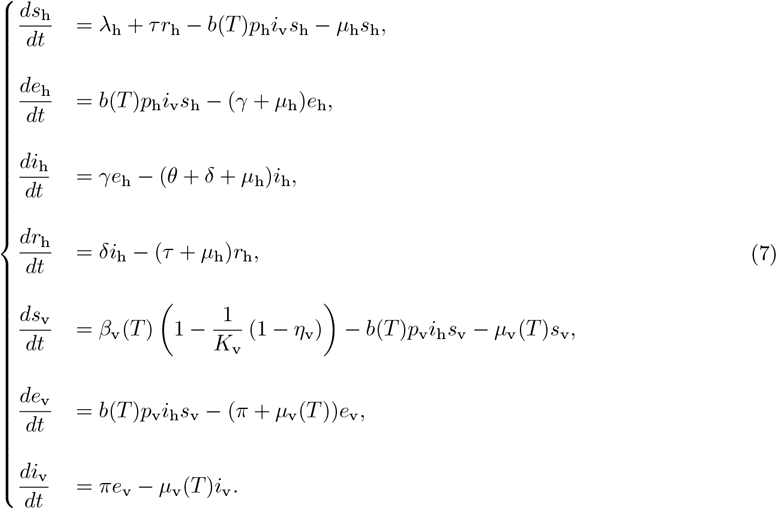

### 3.1 Boundedness and Positivity of solutions

#### 3.1.1 Boundedness

Consider the positively invariant region

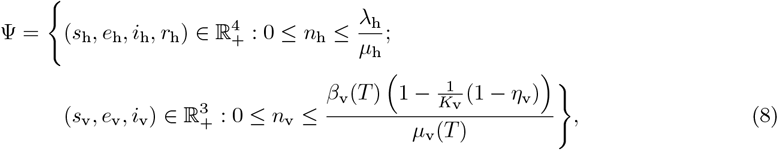

where *n*_h_ = *s*_h_ + *e*_h_ + *i*_h_ + *r*_h_ and *n*_v_ = *s*_v_ + *e*_v_ + *i*_v_.

##### Theorem 1

*All solutions of System* (7) *with initial conditions in* Ψ *remain bounded in* Ψ *for all t* ≥ 0.

*Proof* We consider the human and vector sub-systems separately. Summing the first four equations of System (7) yields

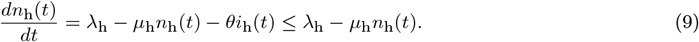

Solving this differential inequality gives

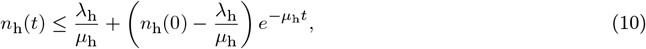

and hence 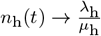 as *t* → ∞. Similarly, summing the vector sub-system equations gives

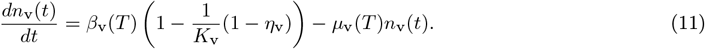

Solving yields

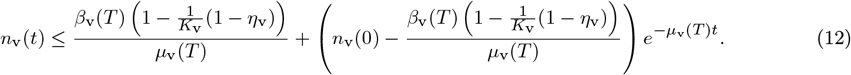

Thus *n*_v_(*t*) remains bounded for all *t* ≥ 0. Therefore, both the human and vector populations are uniformly bounded, and the solution set of System (7) remains in Ψ. □

#### 3.1.2 Positivity of solutions

##### Theorem 2

*If the initial conditions satisfy*

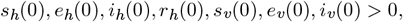

*then the solutions of System* (7) *satisfy*

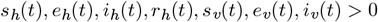

*for all t >* 0.

*Proof* Let

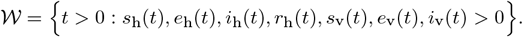

Assume 𝒲 is bounded with supremum *t*_*s*_ *>* 0. By continuity of solutions, *t*_*s*_ exists and is finite only if one of the state variables reaches zero at *t*_*s*_.

Consider the first equation of System (7):

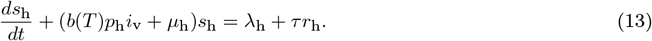

Using the integrating factor

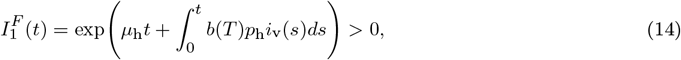

the solution of Equation (13) is

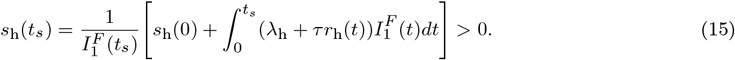

Similarly, applying the same method to the other state variables gives

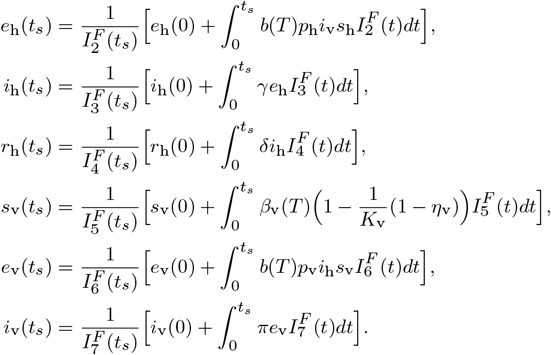

All state variables remain positive on [0, *t*_*s*_], which contradicts the assumption that any variable reaches zero at *t*_*s*_. Therefore, the solutions are positive for all *t >* 0. Thus, System (7) is epidemiologically and mathematically well-posed. □

### 3.2 Basic Reproduction Number, R_**0**_

The Basic reproduction number is the expected number of secondary infections that each case of HAT infection produces during their infectious period within a population of human and tsetse flies that is entirely susceptible [23, 24]. The next generation matrix *FV* ^−1^ is obtained following Van den Driessche and Watmough approach explained in [25] as follows:

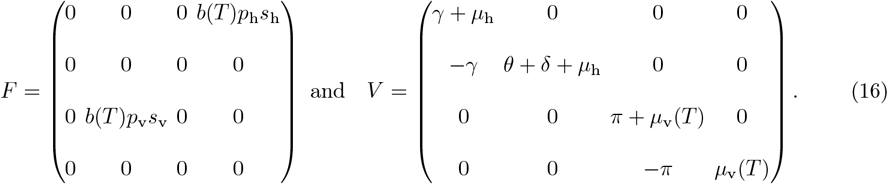

We obtain

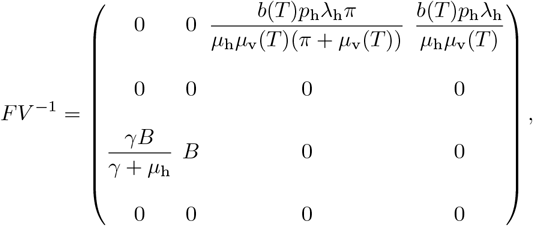

Where 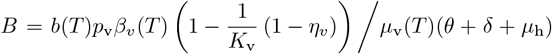. ℜ_0_ is the spectral radius, *ρ*(*FV* ^−1^), therefore

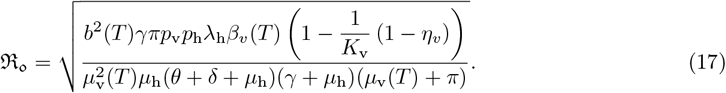

The transmission of the HAT disease occurs when the two populations (the tsetse vector and human populations) are in interplay. Therefore, we can express the basic reproductive number as 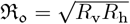 where

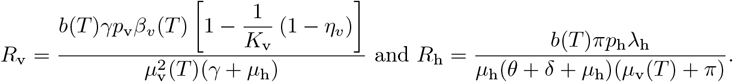

*R*_v_ indicates the number of infections caused by introducing an infected human host into a susceptible tsetse fly population, while *R*_h_ indicates the number of infections in humans caused by an infected tsetse fly introduced into a wholly susceptible human population. Hence, we observe that ℜ_0_ is the geometric mean of *R*_v_ and *R*_h_.

### 3.3 Existence and Stability of Equilibria

The Disease Free Equilibrium (DFE) occurs when there are no HAT infections in the human and tsetse populations [11]. This is obtained as;

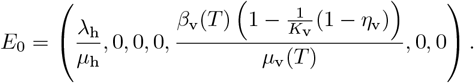

On the other hand, the Endemic Equilibrium (EE) exists when HAT persists in the populations. It is obtained as

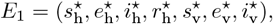

such that

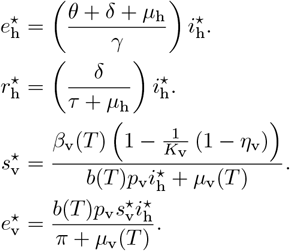

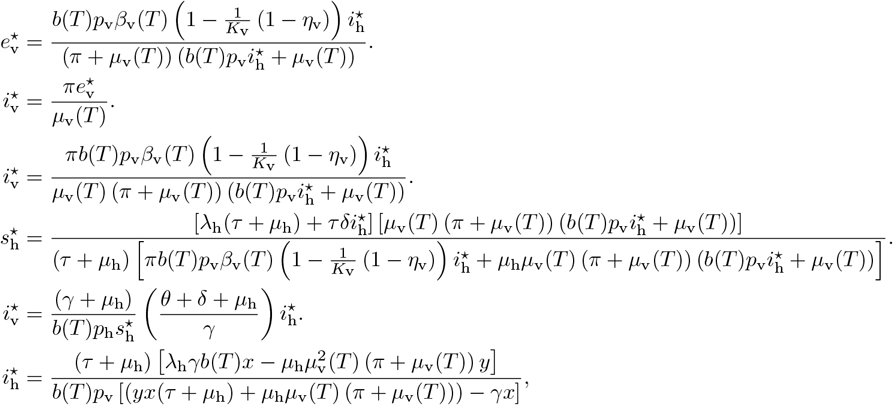

where, 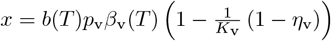 and *y* = (*γ* + *µ*_h_) (*θ* + *δ* + *µ*_h_) .

### 3.4 Stability Analysis of the Equilibria

#### 3.4.1 Local Stability of the DFE

The DFE is locally asymptotically stable if all eigenvalues of the Jacobian matrix evaluated at *E*_0_ have negative real parts [26, page 33].

##### Theorem 3

*The DFE, E*_0_, *is locally asymptotically stable if* ℜ_0_ *<* 1 *[27]*.

*Proof* Consider the submatrix *G* = *F* − *V* corresponding to the infected compartments. Therefore,

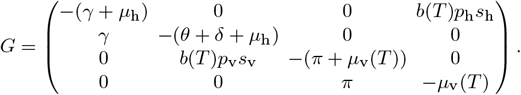

The matrix *G* is a Metzler matrix with non-negative off-diagonal entries. It is stable if *ρ*(*FV* ^−1^) = ℜ_0_ *<* 1, since −*G* = *V* − *F* is a non-singular M-matrix [26]. Hence, all eigenvalues of *G* have negative real parts, and *E*_0_ is locally asymptotically stable. □

### 3.5 Stability Analysis of the Equilibria

#### 3.5.1 Local Stability of the DFE

The DFE point is asymptotically stable if all eigenvalues of the Jacobian matrix of the system have negative real parts [26, page 33]. The Jacobian matrix for System (7) evaluated at *E*_0_ is given by

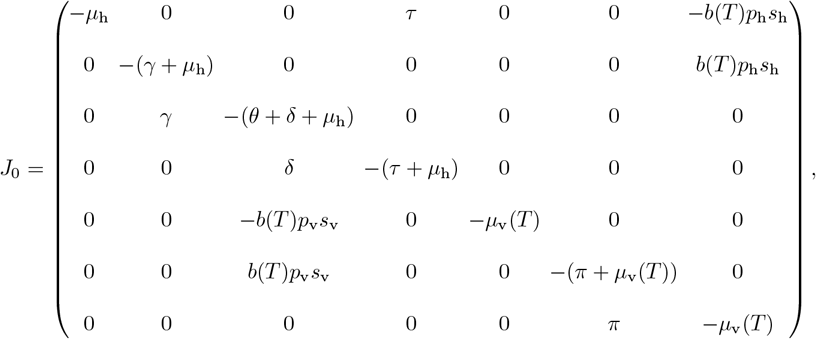

Where

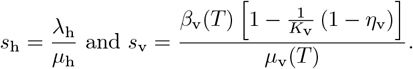

##### Theorem 4

*The DFE point E*_0_ *of the system is locally asymptotically stable if* ℜ_0_ *<* 1 *[27]*.

*Proof E*_0_ is said to be locally asymptotically stable if all eigenvalues of *J*_0_ have negative real parts and unstable if at least one eigenvalue has positive real part. Consider the matrix *G* = *F* − *V*, (a submatrix from *J*_0_).

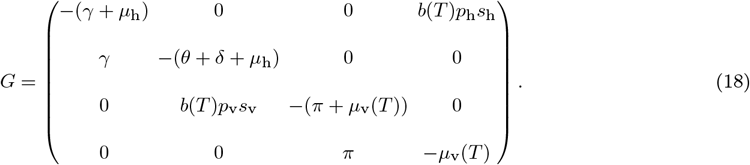

It is clear that the matrix *G* has non negative entries in the off diagonal entries. *G* is therefore a *Metzler matrix*. The matrix *G* is stable whenever *ρ*(*FV* ^−1^) = ℜ_0_ *<* 1. This follows from the fact that the matrix −*G* = *V* − *F* is a non-singular M-matrix for ℜ_0_ *<* 1 (See the proof of Theorem 2 in Van den Driessche and Watmough [26], pages 33-34). The spectral radius, *ρ*(*G*) *<* 0 if ℜ_0_ *<* 1. This implies that all eigenvalues of *G* have negative real parts. This completes the proof. □

#### 3.5.2 Local Stability of the Endemic Equilibrium

##### Theorem 5

*The endemic equilibrium point*, 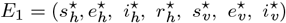 *is locally asymptotically stable in the defined region* Ψ *if* ℜ_0_ *>* 1 *and unstable otherwise*.

*Proof* Given that

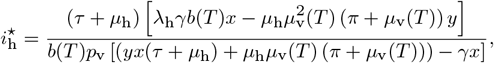

it implies that, at the Endemic Equilibrium point, the disease persists within the human population and therefore 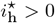 (see [11]). Therefore,

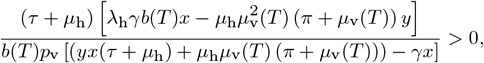

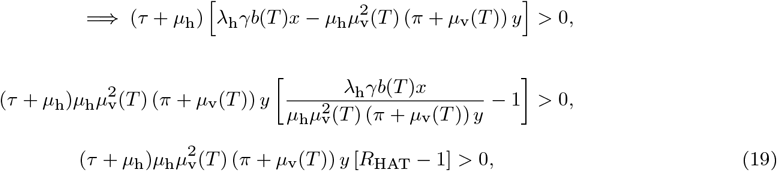

where

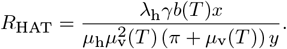

For the inequality in Equation (19) to hold, *R*<SUB>HAT </SUB>– 1 > 0, which follows that *R*<SUB>HAT </SUB>>1. Notice that 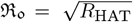. This implies that ℜ_0_ *>* 1. □

#### 3.5.3 Global Stability of the DFE

##### Theorem 6

*The DFE point E*_0_ *of System* (7) *is globally asymptotically stable in the region* Ψ *whenever* ℜ_0_ *<* 1 *[28, page 9]*.

*Proof* Let ℒ (*t*) be a Lyapunov function given by

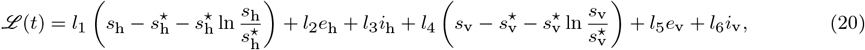

where *l*_1_, *l*_2_, *l*_3_, *l*_4_, *l*_5_ and *l*_6_ are positive constants which are to be determined and

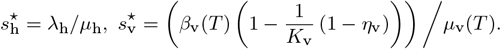

Observe that the function *r*_h_(*t*) does not take part in the function ℒ and ignored in the substitutions below since *r*_h_(*t*) → 0 at any time when the DFE for *s*_h_, *e*_h_, *i*_h_ is globally stable and this suffices for the whole system to be globally stable. It can easily be shown that ℒ (*t*) = 0 at the DFE point. Now, we need to confirm that ℒ (*t*) *>* 0 for all

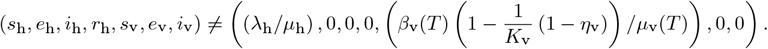

It suffices to show that at a point different from the DFE,

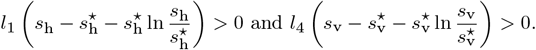

Let us consider,

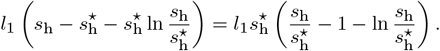

Let *h*(*a*) = *a* − 1 − ln *a* where 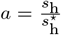. Clearly, 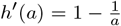 and 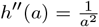. Therefore, the function *h*(*a*) has a minimum value 0 which occurs when *a* = 1. This implies that *h*(*a*) *>* 0, ∀*a >* 0, *a* ≠ 1. It follows that the first term and the fourth terms of Equatio n (20) a re pos itive and thus ℒ *>* 0 for all 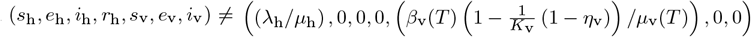. ℒ is radially unbounded since *h*(*a*) → ∞ when ∥ *a* ∥→ ∞. Differentiating Equation (20) with respect to time (*t*)

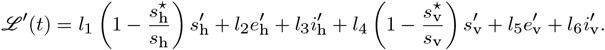

Substituting the derivatives of the state variables using the right hand side of the respective equations in the system, we obtain

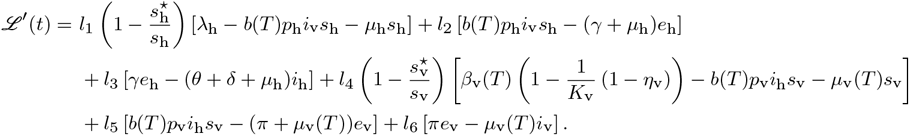

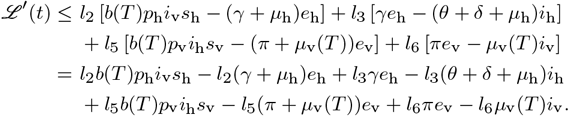

Grouping the terms according to the state variables *i*_h_, *e*_h_, *i*_h_, *e*_v_, we obtain

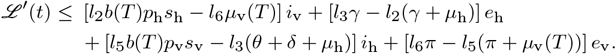

At the DFE, *s*_h_ = *λ*_h_*/µ*_h_ and 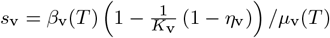. It implies that, ℒ^*′*^(*t*) becomes,

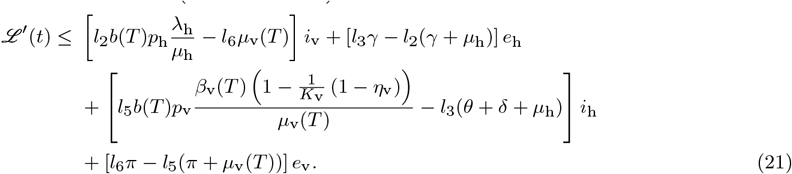

Equating the coefficients of *i*_*v*_ and *i*_*h*_ in Equation (21) to zero,

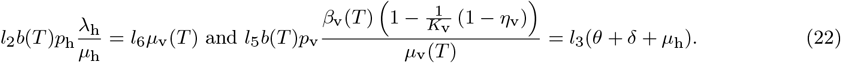

From Equation (22), we make choices

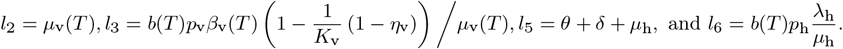

Substituting *l*_2_, *l*_3_, *l*_5_, and *l*_6_ into Equation (21) gives

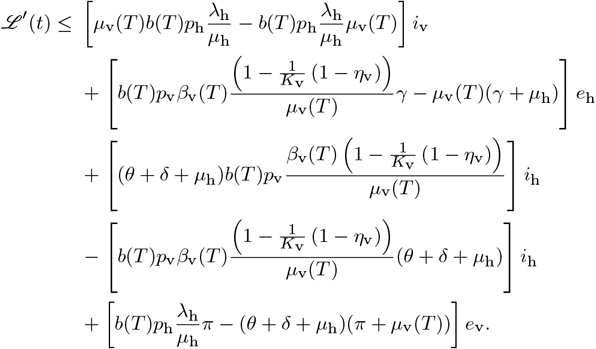

The terms which are multiplied by *i*_*v*_ and *i*_*h*_ vanish which gives

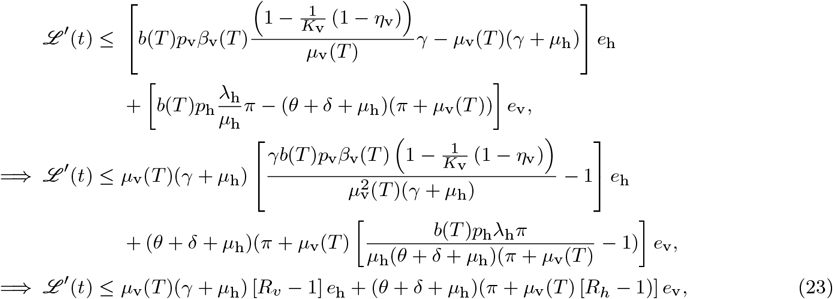

where

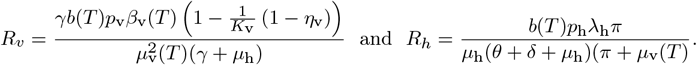

From Equation (23), we note that ℒ^*′*^(*t*) *<* 0 if *R*_*h*_, *R*_*v*_ *<* 1 and ℒ^*′*^(*t*) = 0 if and only if *e*_v_, *e*_h_ = 0. It therefore follows that, the unit set

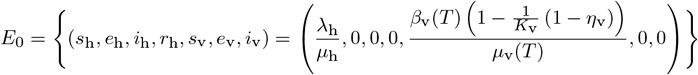

is the only largest compact invariant set in the region Ψ defined in Equation (8). Thus, from *La Salle’s Invariance principle*, it can be concluded that the DFE *E*_0_ is globally asymptotically stable in the region Ψ provided ℜ _0_ *<* 1. □

### 3.6 Bifurcation Analysis

We determine the direction of the bifurcation for System (7) using the *center manifold theory* as discussed in [29] by setting the basic reproduction number, ℜ_0_ to one and making the transmission rate, *p*_h_, our bifurcation parameter in ℜ_0_ [27]. We can therefore express the bifurcation parameter 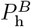 (the parameter *p*_h_ at ℜ_0_ = 1) as follows;

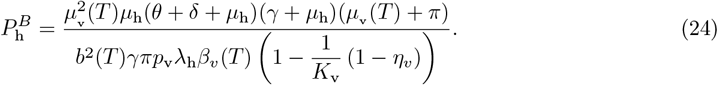

The state variables are then re-written as

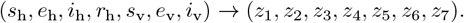

*Z* = (*z*_1_, *z*_2_, *z*_3_, *z*_4_, *z*_5_, *z*_6_, *z*_7_)^*T*^ is a vector which now produces a new modified system of the form

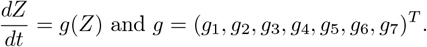

The expanded system is

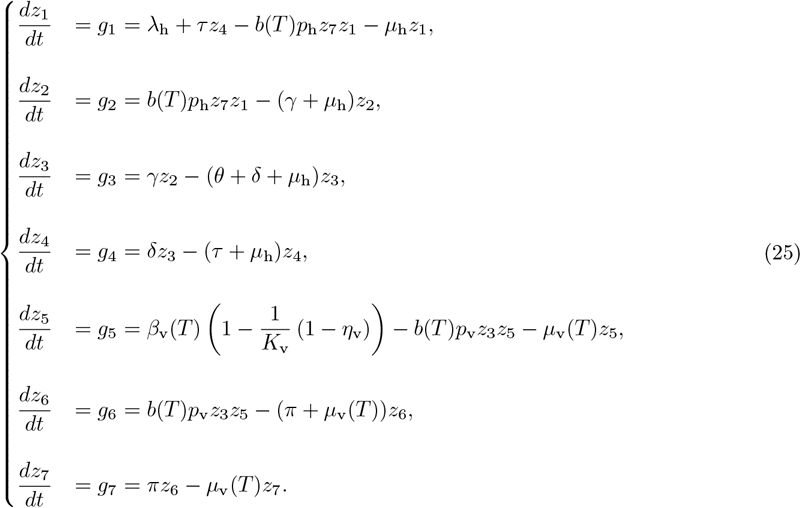

The Jacobian matrix for System (25) evaluated at DFE is given by

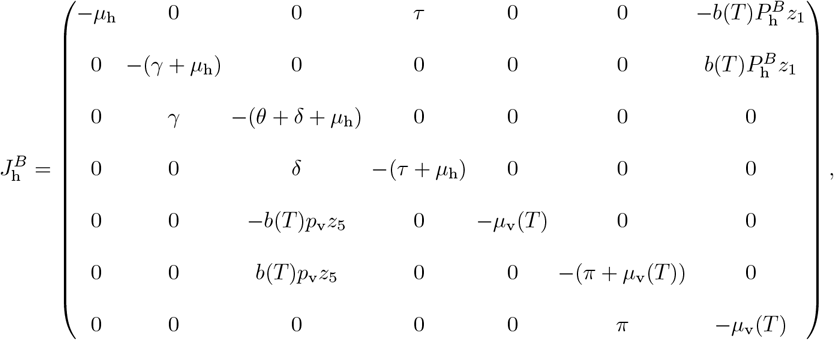

Where

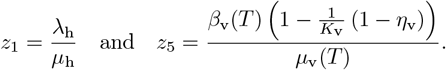

Since, the Jacobian matrix 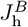 has a simple zero as one of its eigenvalues and all the rest having negative real parts, we satisfy the condition to employ the center manifold theory. Next, we determine the left and right eigenvectors. The right eigenvector, **X**, of the matrix is one that satisfies 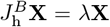. Therefore, it can be computed by solving

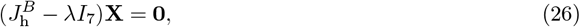

where *λ* is an eigenvalue, **X** is a column vector and *I*_7_ is the 7 × 7 identity matrix. Solving Equation (26), we obtain the right eigenvector associated to *λ* = 0 as **X** = (*x*_1_, *x*_2_, *x*_3_, *x*_4_, *x*_5_, *x*_6_, *x*_7_)^*T*^, where

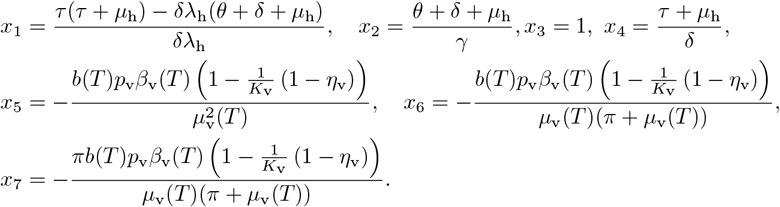

The left eigenvector, **Y**, of the matrix 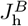 satisfies 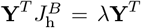. Therefore, it can be computed by solving

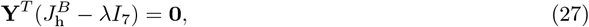

where **Y** is a column vector. Solving Equation (27), we obtain the left eigenvector associated to *λ* = 0 as **Y** = (*y*_1_, *y*_2_, *y*_3_, *y*_4_, *y*_5_, *y*_6_, *y*_7_)^*T*^, where

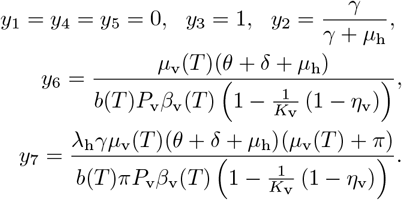

The second partial derivatives of *g* at DFE are

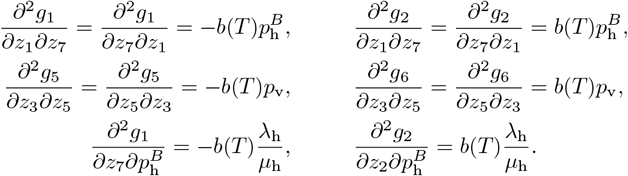

The coefficients *a* and *b* are defined by

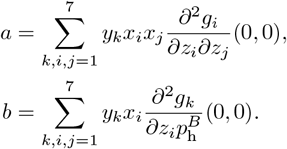

This yields

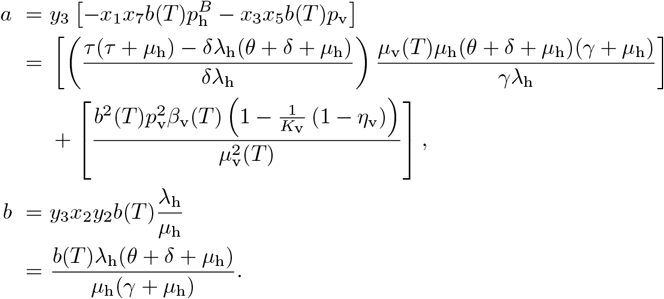

Clearly, *b* is always positive and therefore, the dynamics around the DFE can be determined based on the sign of *a*:

- *a* is positive if 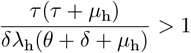. This implies a backward bifurcation.
- *a* is negative if 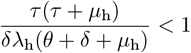. This implies a forward bifurcation.

### 3.7 Optimal Control Analysis

We introduce four time-dependent control functions, *c*_1_(*t*), *c*_2_(*t*), *c*_3_(*t*) and *c*_4_(*t*) into System (7). The control functions *c*_*j*_(*t*); *j* = 1, 2, 3, 4 can take values ranging from 0 to 1, that is, 0 ≤ *c*_*j*_(*t*) ≤ 1, *t* ∈ [0, *T*]. For convenience, we shall write *c*_*j*_ for *c*_*j*_(*t*). The controls 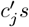 *s* indicate the level of effectiveness of the HAT control strategies and are explicitly defined as;

- *c*_1_ represents the level of *personal protection* by wearing gloves, long-sleeved shirts and pants during day-time to minimise interactions between the human and tsetse flies.
- *c*_2_ indicates *treatment* as a strategy to control HAT infections.
- *c*_3_ shows the use of *bed nets* to minimize the contacts between the humans and the tsetse flies.
- *c*_4_ represents the efforts of *insecticide spray* in order to reduce the breeding sites of tsetse flies.

The modified system in which the controls have been incorporated is now given by

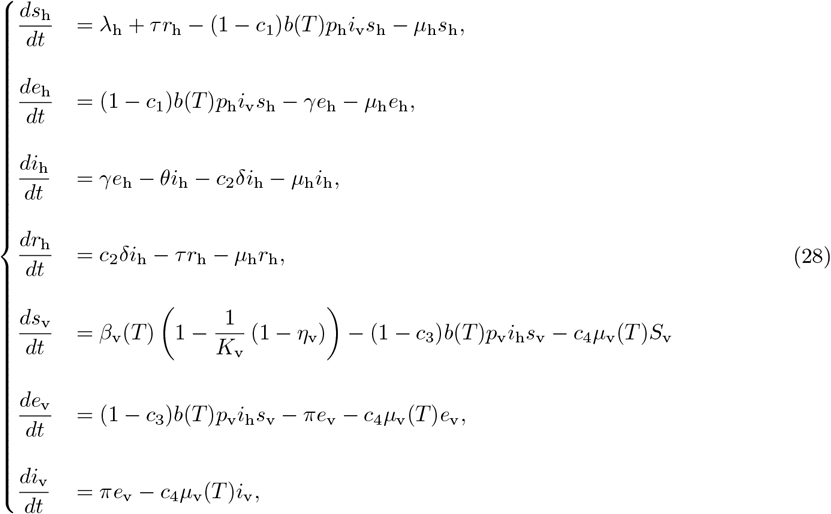

subject to *s*_h_ ≥ 0, *e*_h_ ≥ 0, *i*_h_ ≥ 0, *r*_h_ ≥ 0, *s*_v_ ≥ 0, *e*_v_ ≥ 0 and *i*_v_ ≥ 0. The control variables *c*_*j*_(*t*) are subject to the state variables *s*_h_, *e*_h_, *i*_h_, *r*_h_, *s*_v_, *e*_v_, *i*_v_ which are measured and bounded by

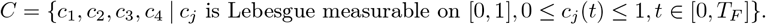

We define the objective function for our optimal control problem as

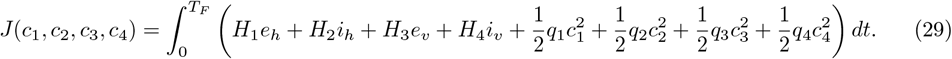

The quantities *H*_1_ and *H*_2_ account for the cost of minimising the exposed and infected humans, respectively, whereas *H*_3_ and *H*_4_ represent the respective cost of minimising the exposed and infectious tsetse flies. The terms *H*_1_*e*_*h*_, *H*_2_*i*_*h*_, *H*_3_*e*_*v*_, and *H*_4_*i*_*v*_ are linear. The costs of enforcing personal protection, treatment, sleeping under tsetse fly bed nets and tsetse flies spray activities are given by the quadratic terms 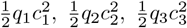, and 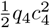, respectively. The period of control intervention is denoted by *T*_*F*_ . Therefore, we minimise the objective function over the interval [0, *T*_*F*_].

We use the Pontryagin’s maximum principle to solve the optimal control problem and obtain the necessary conditions. From Equation (29), the Lagrangian *L* is given by

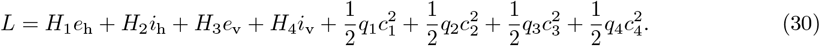

We define the Hamiltonian, *H*, as

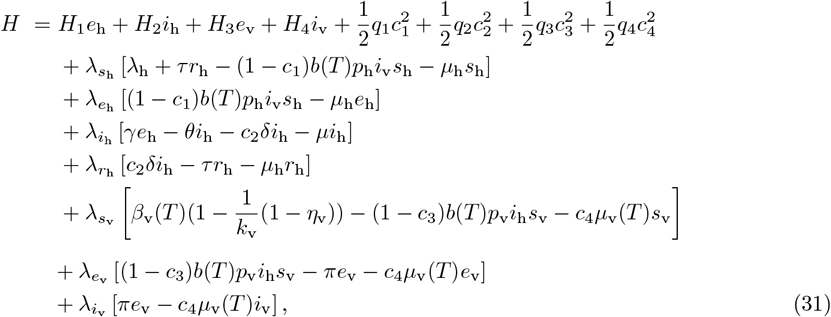

where *λ*_*k*_ are the adjoint variables. We then obtain the system of differential equations of the adjoint variables by using 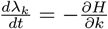, where *k* ∈ (*s*_h_, *e*_h_, *i*_h_, *r*_h_, *s*_v_, *e*_v_, *i*_v_), such that

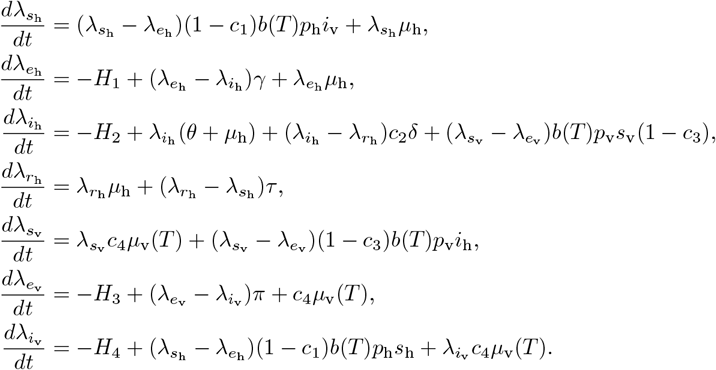

#### Theorem 7

*Given the optimal controls* 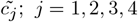 *and the solutions of the state variables of System* (28), *which minimise the objective functional J* (*c*_1_, *c*_2_, *c*_3_, *c*_4_) *defined in Equation* (29) *over the interior of the control set C, there exists adjoint variables* 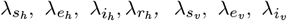 *satisfying the system*

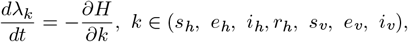

*and with transversality conditions λ*_*k*_ (*T<SUB>F </SUB>*) = 0 *and the optimality conditions given by* 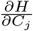, *with the controls given by*

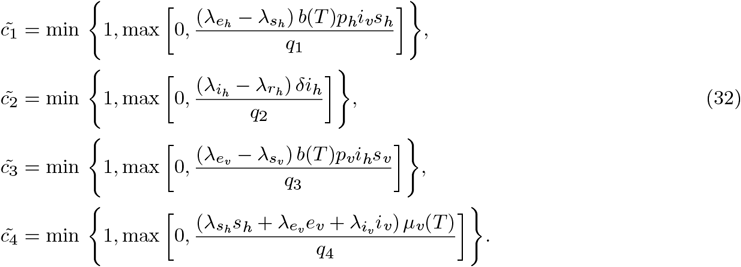

*Proof* The partial derivatives of the Hamiltonian with respect to the controls are

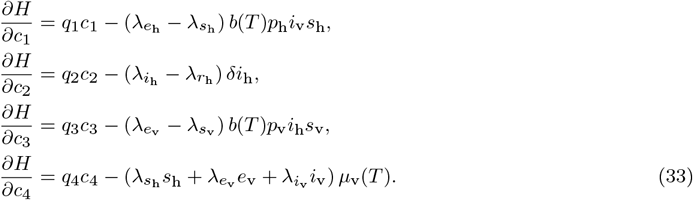

But, the optimality conditions are given as

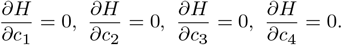

Applying the above conditions to solve Equation (**??**) yields

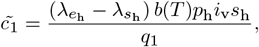

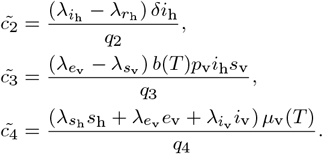

## 4 Numerical Results

This section presents numerical simulations of System (28). The model parameters were estimated using the least-squares method, and the resulting values summarized in Table 2. The initial conditions were set as

**Table 2.** Estimated values of the model parameters.

| Parameter | Value | Source |
| --- | --- | --- |
| $\lambda_h$ | 0.095 | Estimated |
| $\beta_v$ | 0.07 | [11] |
| $b$ | 0.7 | [11] |
| $p_h$ | 0.02755918 | Estimated |
| $p_v$ | 0.065 | [11] |
| $\mu_h$ | 0.000284422 | Estimated |
| $\mu_v$ | 0.034 | [11] |
| $\gamma$ | 0.002756617 | Estimated |
| $\theta$ | 0.01644727 | Estimated |
| $\delta$ | 0.60 | Estimated |
| $\tau$ | 0.041062 | Estimated |
| $\pi$ | 0.001 | [11] |
| $K_v$ | 10000 | Assumed |
| $\eta_v$ | 0.05 | Assumed |

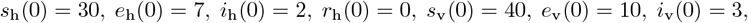

while the weighting and balancing parameters were chosen as

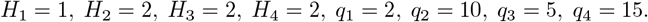

Simulations were carried out over a 120-day period using the forward–backward sweep algorithm [30], coupled with a fourth-order Runge–Kutta scheme implemented in `OCTAVE`.

### 4.1 Intervention 1: Using Single Control strategy

Figures A.1a–A.1f and A.2a–A.2f (see supplementary material) show the effects of single control strategies on the dynamics of HAT in human and tsetse fly populations. Insecticide spraying leads to a significant reduction in infected vectors (see Figure A.1b), while bed net usage results in a modest decrease in vector infections (see Figure A.1d). Treatment effectively reduces the infected human population (see Figure A.2a) but not the tsetse fly population. The results indicate that a single control strategy is insufficient for eliminating HAT in both populations.

### 4.2 Intervention 2: Using two control strategies

Figures A.3a–A.3f, A.4a–A.4f, and A.5a–A.5f (see supplementary material) present the simulation results for combinations of two control strategies. The combined use of bed nets and insecticide spraying reduces the infectious vector population (see Figure A.3b). Similarly, treatment coupled with insecticide spraying lowers both infected humans and vectors (see Figures A.3c and A.3d), although the reduction in infected humans were marginal. The combination of personal protection and insecticide spraying also decreases infections in both populations (see Figures A.4c and A.4d).

### 4.3 Intervention 3: Using three control strategies

Figures A.6a–A.6f and A.7a–A.7f (see supplementary material) show that various combinations of three control strategies substantially reduce infections in both human and tsetse fly populations. In particular, the combined use of treatment, personal protection, and insecticide spraying effectively reduce transmission and can potentially eliminate HAT (see Figures A.6c and A.6d). However, not all three-control combinations are equally effective. For instance, the combination of treatment, personal protection, and bed nets fails to eliminate infection in the vector population (see Figure A.7d).

### 4.4 Intervention 4: Using all control strategies

Figures A.8a–A.8c (see supplementary material) demonstrate that the simultaneous implementation of personal protection, treatment, bed net usage, and insecticide spraying results in a substantial reduction of infections in both human and tsetse fly populations. This combined strategy is the most effective across all compartments and represents the optimal intervention for eliminating HAT from both populations.

### 4.5 Temperature Analysis

Using the mean temperature time series data from [31] for Central African Republic which is used for our case study since it is listed among the countries at the highest risk of acquiring HAT disease according to [32]. The total mean temperatures over the years 2010 to 2025 were calculated which were used to calculate the average temperatures over the same period for each month, and the values are shown in Table 3.

**Table 3.** The total average monthly temperature values in ^*°*^ C over the years 2010 to 2025: T stands for Temperatures and M stands for Months of the year.

| M | Jan | Feb | Mar | Apr | May | Jun | Jul | Aug | Sep | Oct | Nov | Dec |
| --- | --- | --- | --- | --- | --- | --- | --- | --- | --- | --- | --- | --- |
| T | 24.62 | 26.93 | 27.53 | 27.41 | 26.50 | 25.24 | 24.43 | 24.07 | 24.36 | 24.76 | 24.94 | 24.39 |

Figure 2, shows the line graph of the average monthly temperatures over the years 2010 to 2025.

**Fig. 2.**
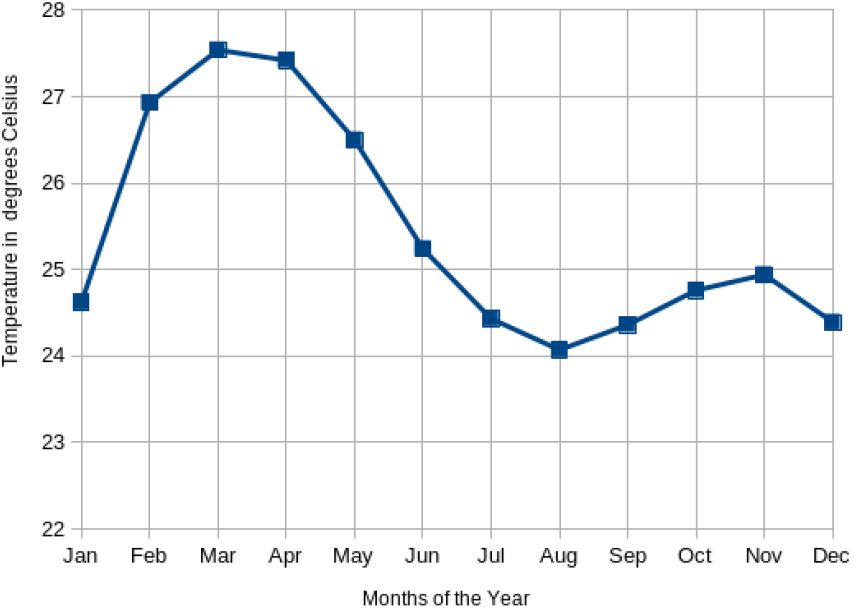
The average mean monthly temperature for the Central African Republic

Mean temperature is a key determinant of tsetse fly survival and abundance [8]. Tsetse flies cannot survive at temperature extremes below 16^*°*^C or above 32^*°*^C [10, 19]. Moreover, their reproduction, development, and mortality rates are strongly temperature-dependent [33]. Tsetse fly abundance increases between 21^*°*^C and 25^*°*^C but declines at higher temperatures, particularly between 26^*°*^C and 30^*°*^C. These findings suggest that disease control measures should be intensified during periods when temperatures favor tsetse fly proliferation and can be relaxed during hotter periods when vector populations naturally decline. Since high temperatures reduce tsetse fly survival, HAT transmission is correspondingly reduced. Consequently, incorporating seasonal temperature variation into disease models enhances understanding of HAT transmission dynamics and supports the development of more effective, seasonally targeted control strategies.

### 4.6 Estimation of Temperature-Dependent Parameters

To capture the effect of temperature on HAT transmission, the biting rate *b*(*T*), transmission probability *β*_*v*_(*T*), and vector mortality rate *µ*_*v*_(*T*) are modeled as temperature-dependent functions using biologically validated formulations for vector-borne diseases.

#### 4.6.1 Biting Rate *b*(*T*)

The biting rate is modeled using a Brière-type function, which increases with temperature up to an optimal range and vanishes at thermal extremes:

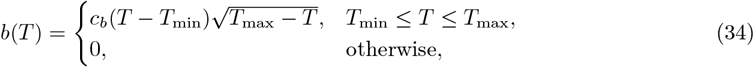

where *T*_min_ = 23^*°*^C and *T*_max_ = 30^*°*^C reflect observed monthly temperature bounds. The constant *c*_*b*_ is estimated using the mean biting rate *b*_mean_ = 0.7 at 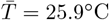.

#### 4.6.2 Transmission Probability *β*_*v*_(*T*)

The transmission probability per bite is similarly described by a Brière function:

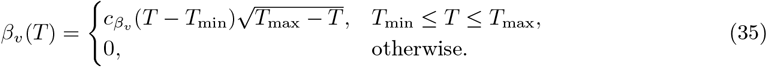

Using *β*_*v*,mean_ = 0.07 at 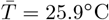, the parameter 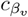 is determined accordingly.

#### 4.6.3 Vector Mortality Rate *µ*_*v*_(*T*)

Vector mortality is assumed to increase away from an optimal temperature and is modeled by a quadratic function:

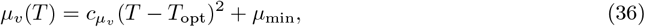

where *T*_opt_ = 26^*°*^C and *µ*_min_ = 0.01. The coefficient 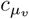 is estimated using the observed mean mortality *µ*_*v*,mean_ = 0.034.

#### 4.6.4 Monthly Parameter Evaluation

Mean monthly temperatures are substituted into the above expressions to compute discrete monthly values of *b*(*T*), *β*_*v*_(*T*), and *µ*_*v*_(*T*), which are used in numerical simulations. The resulting parameter values are summarized in Table 4 and their seasonal variation is illustrated in Figures 3 and 4.

**Table 4.**
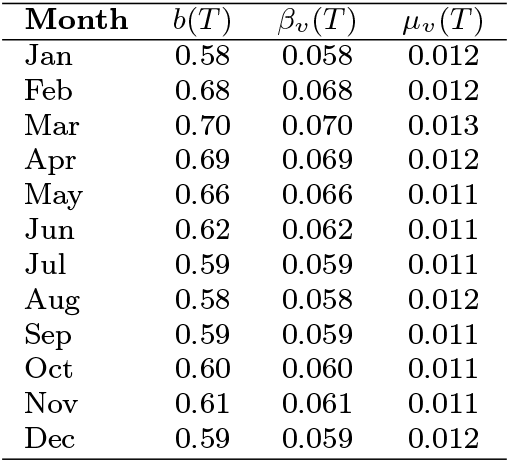
Monthly temperature-dependent parameter values for *b*(*T*), *β*_*v*_(*T*), and *µ*_*v*_(*T*).

**Fig. 3.**
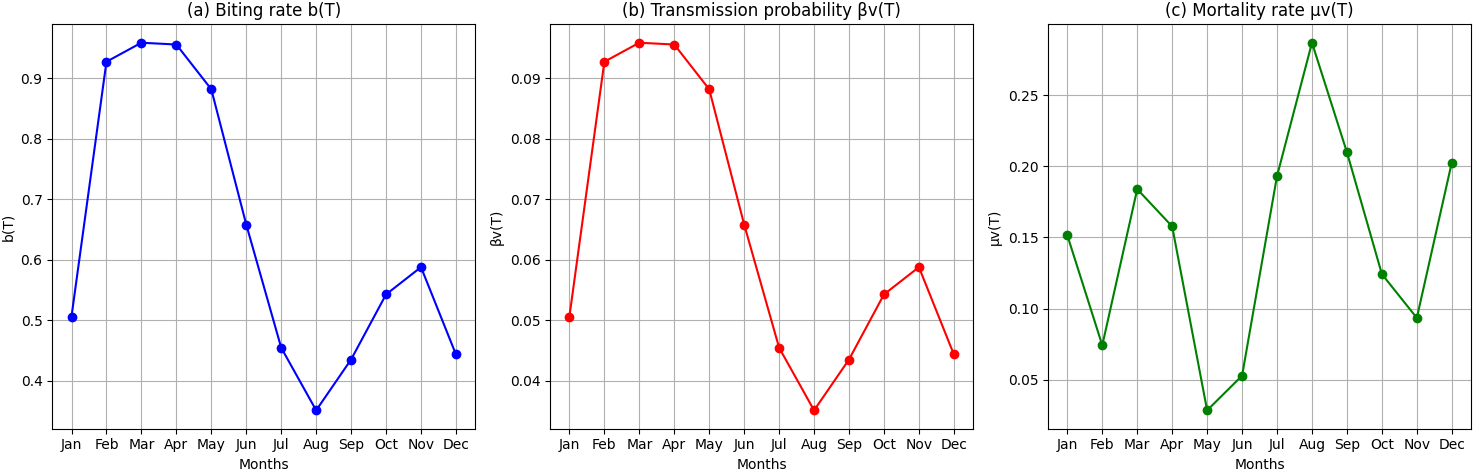
Temperature-dependent parameters. (a) Biting rate *b*(*T*); (b) Transmission probability *β*_*v*_(*T*); (c) Mortality rate *µ*_*v*_(*T*). Each point represents the parameter value calculated for the mean monthly temperature (Jan–Dec).

**Fig. 4.**
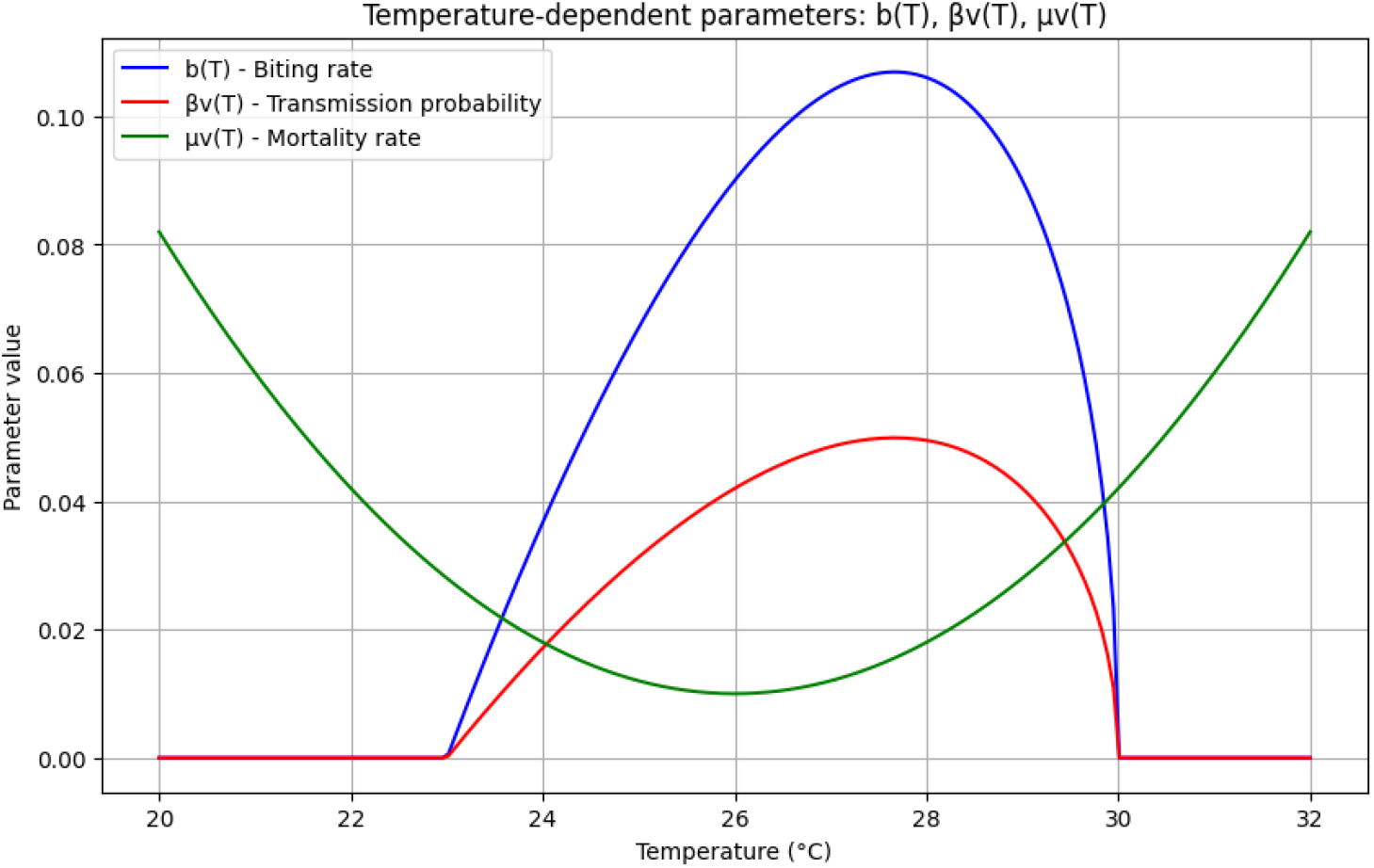
Plots of the temperature-dependent parameters.

Temperature strongly influences HAT transmission dynamics. As shown in Table 4 and Figures 3 and 4, the biting rate *b*(*T*) and transmission probability *β*_*v*_(*T*) peak during moderate temperatures (February–April), indicating optimal conditions for tsetse activity and parasite transmission. Both parameters decline during hotter months, reflecting reduced vector efficiency. Conversely, the mortality rate *µ*_*v*_(*T*) is lowest at moderate temperatures and increases at thermal extremes, reducing vector survival. Thus, HAT transmission potential is highest within a narrow optimal temperature range, highlighting the importance of seasonally targeted control interventions.

## 5 Conclusion

This paper advances the mathematical epidemiology of Human African Trypanosomiasis by explicitly embedding temperature-dependent vector biology within a coupled human–tsetse transmission framework. By allowing key entomological parameters, biting rate, recruitment, and mortality, to vary with ambient temperature, the model establishes a direct mechanistic pathway through which environmental variability influences disease persistence, elimination thresholds, and control effectiveness. The explicit derivation of the basic reproduction number reveals that temperature acts nonlinearly on transmission potential, increasing or reducing infection risk through its simultaneous effects on vector survival, development, and host–vector contact rates. Analytical results demonstrate that the disease-free equilibrium is globally asymptotically stable when (*R*_0_ *<* 1), ensuring elimination under sufficiently unfavorable thermal and epidemiological conditions, while a unique endemic equilibrium emerges and remains locally stable when (*R*_0_ *>* 1). The identification of a forward transcritical bifurcation at the threshold (*R*_0_ = 1) confirms the absence of backward bifurcation in the baseline model, thereby supporting the use of (*R*_0_) as a reliable target for control and elimination strategies. These results provide a mathematically robust foundation for evaluating intervention thresholds under climate-sensitive transmission dynamics. Beyond its theoretical contributions, the model offers important applied insights. Seasonal and longterm temperature variation can shift transmission intensity even in the absence of changes in human behavior or intervention coverage, implying that static control policies may be suboptimal. Incorporating temperature-driven dynamics into surveillance and intervention planning enables more efficient, seasonally targeted control strategies, particularly in regions approaching elimination where small environmental shifts may determine resurgence or fade-out. However, the model assumes homogeneous, deterministic dynamics and relies on seasonal mean temperature, limiting its ability to capture spatial heterogeneity, climatic extremes, and short-term variability. Animal reservoirs and other environmental drivers such as rainfall and humidity are not explicitly included, restricting applicability in zoonotic and ecologically diverse settings. The authors, are in the process of developing spatially explicit, stochastic, and data-calibrated models that integrate high-resolution climate data and reservoir dynamics to improve predictive and policy relevance.

## Supporting information

HAT

## Author contributions

AO: investigation, validation, mathematical formal analysis, parameter estimation and writing original draft, review and editing, NKDOO: conceptualization, model formulation, supervision, investigation, validation, mathematical formal analysis, writing, review and editing final paper. All authors read and approved the final manuscript.

## Data Availability declaration

The data used is stated and provided in the paper.

