## Supplementary material for "Controlling the Transmission Dynamics of HAT Incorporating Impacts of Temperature": HAT

### Numerical Simulations

Simulation graphs for Infectious human and vector populations.

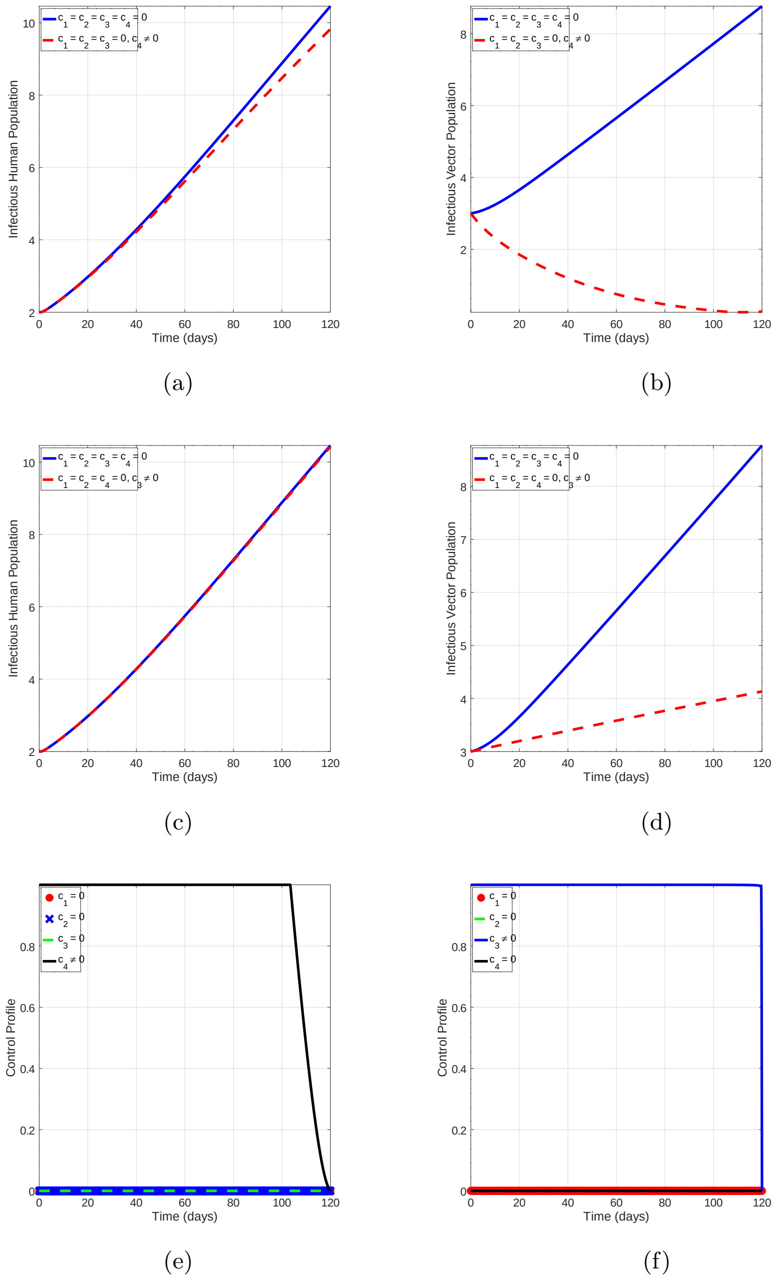


**Fig. A.1**: Simulations of the model (28) showing the efforts of; insecticide spray on the infectious human population (a) and infectious vector population (b). (c) and (d) show the efforts of sleeping under bed nets on the infectious human population and infectious vector populations, respectively. Graphs (e) and (f) show control profiles for the corresponding strategies.

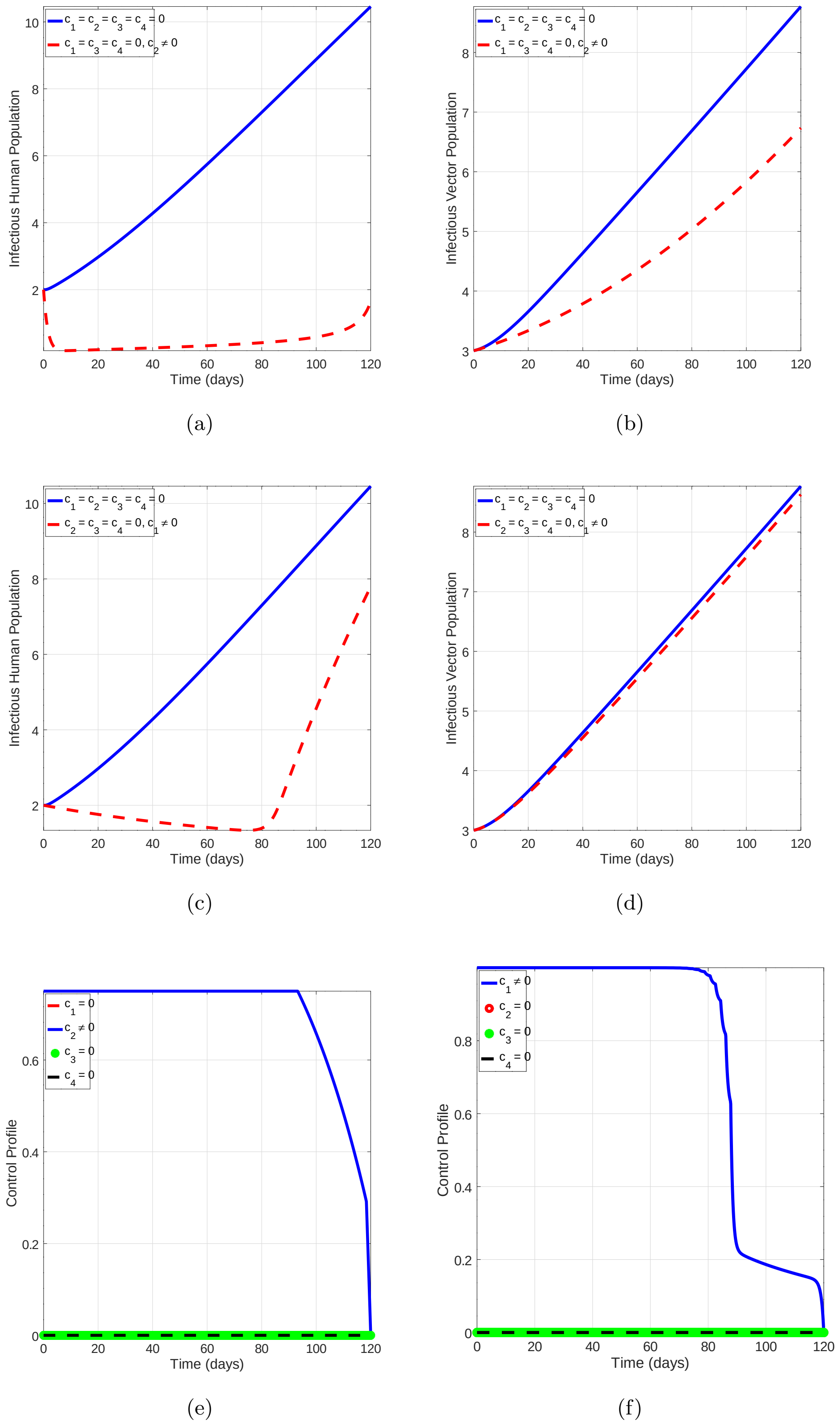


**Fig. A.2**: Simulations of the model (28) showing the efforts of; treatment on (a) the infectious human population and (b) infectious vector populations. Plots (c) and (d) show the efforts of personal protection on the infectious human population and infectious vector populations, respectively. Graphs (e) and (f) show control profiles for the corresponding strategies.

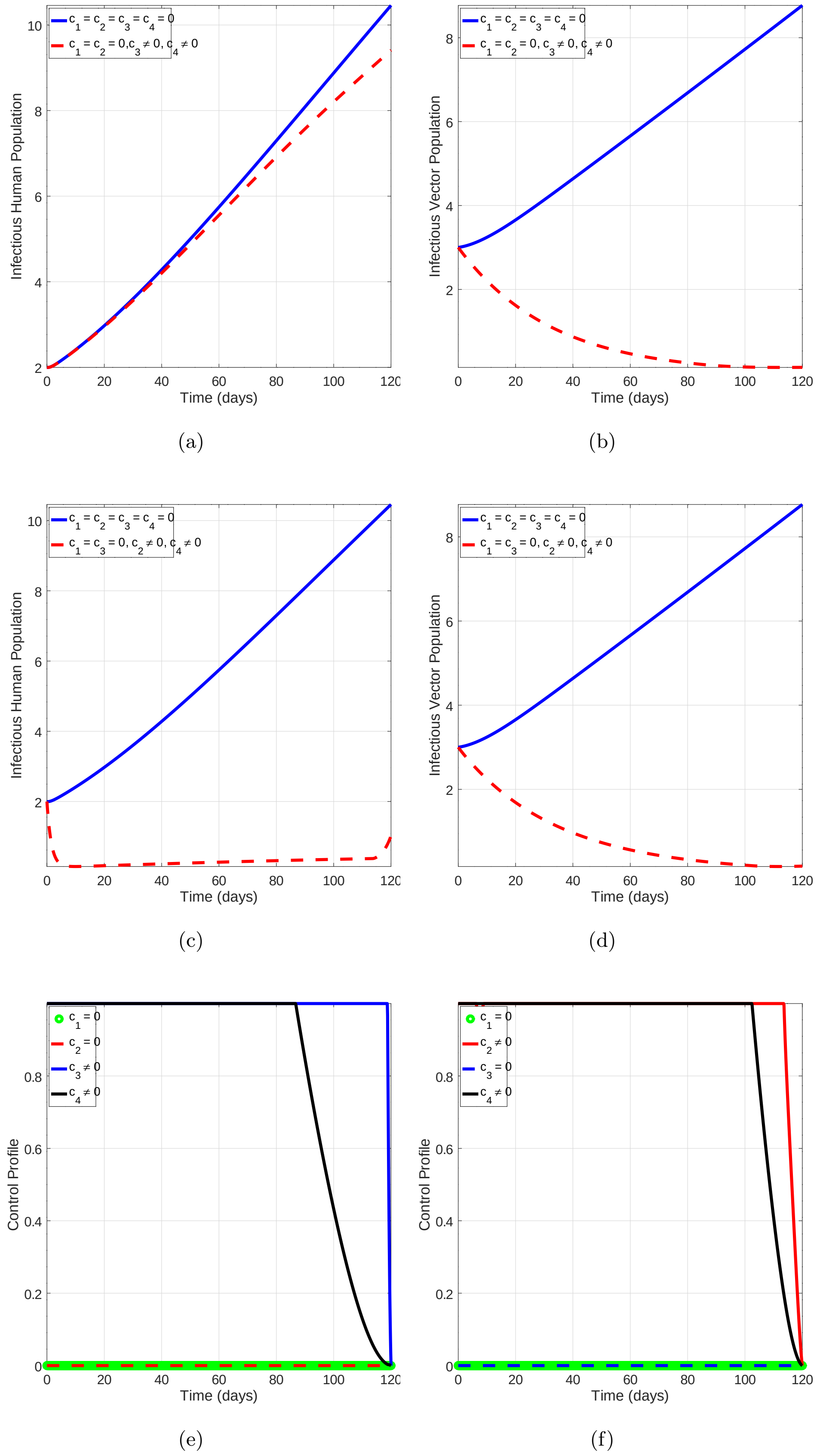


**Fig. A.3**: Simulations of the model (28) showing the efforts of intervention using two controls. (a) and (b) show the efforts of sleeping under bed nets and use of insecticide spray on the infectious human population and infectious vector populations respectively. (c) and (d) show the combined strategy of treatment and insecticide spray on the infectious human and vector populations, respectively. Graphs (e) and (f) show control profiles for the corresponding strategies.

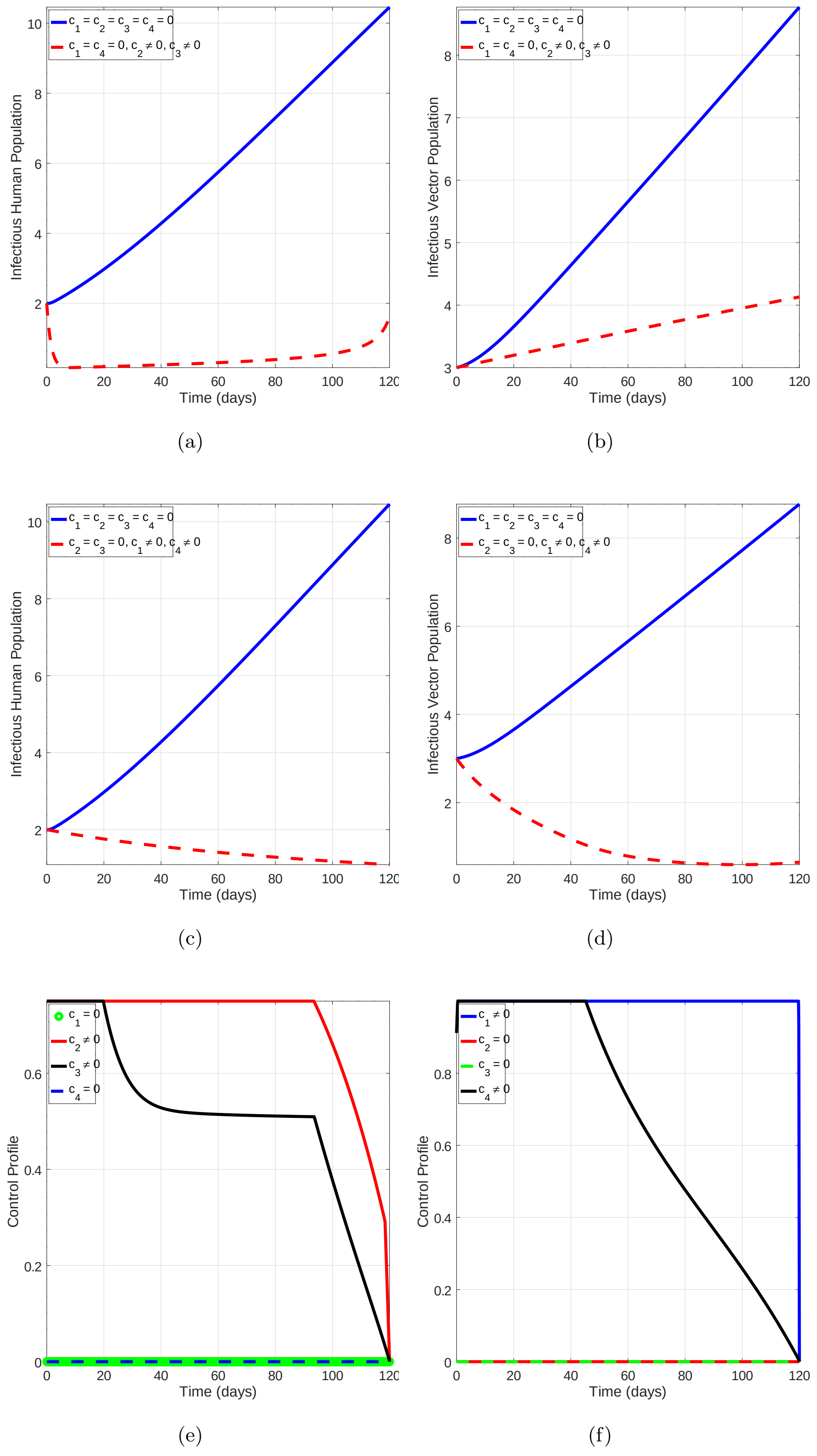


**Fig. A.4**: Simulations of the model (28) showing the efforts of intervention using two controls. (a) and (b) show the efforts of both treatment and bed nets on the infectious human population and infectious vector populations, respectively. (c) and (d) show the combined strategy of using personal protection gadgets with insecticide spray on the infectious human population and infectious vector populations, respectively. Graphs (e)and (f) show control profiles for the corresponding control strategies.

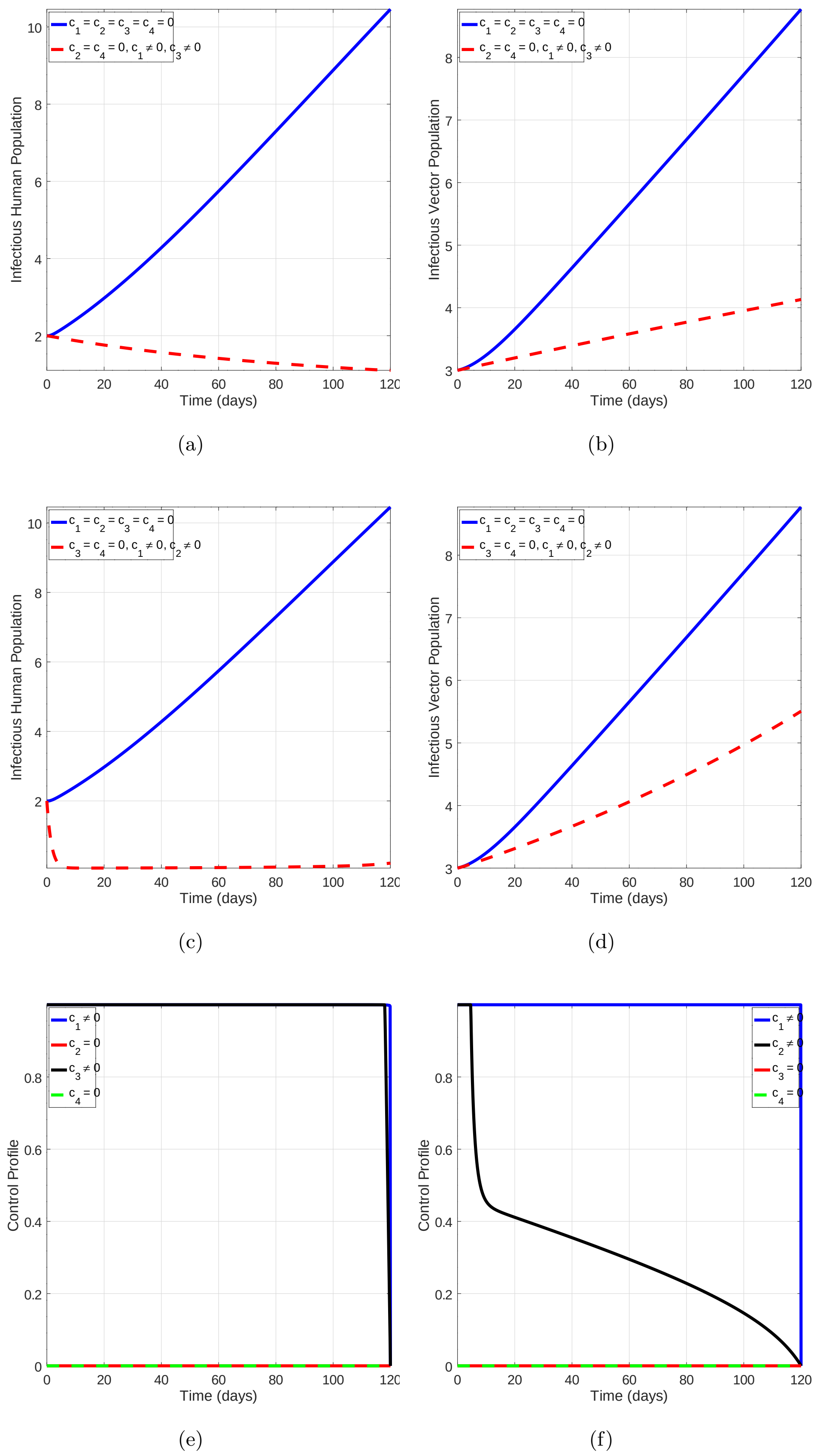


**Fig. A.5**: Simulations of the model (28) showing the efforts of intervention using two controls. (a) and (b) show the efforts of use of both personal protection and bed nets on the infectious human population and infectious vector populations respectively. (c) and (d) show the efforts of using personal protection together with treatment on the infectious human population and infectious vector populations, respectively. Graphs (e) and (f) show control profiles for the corresponding control strategies.

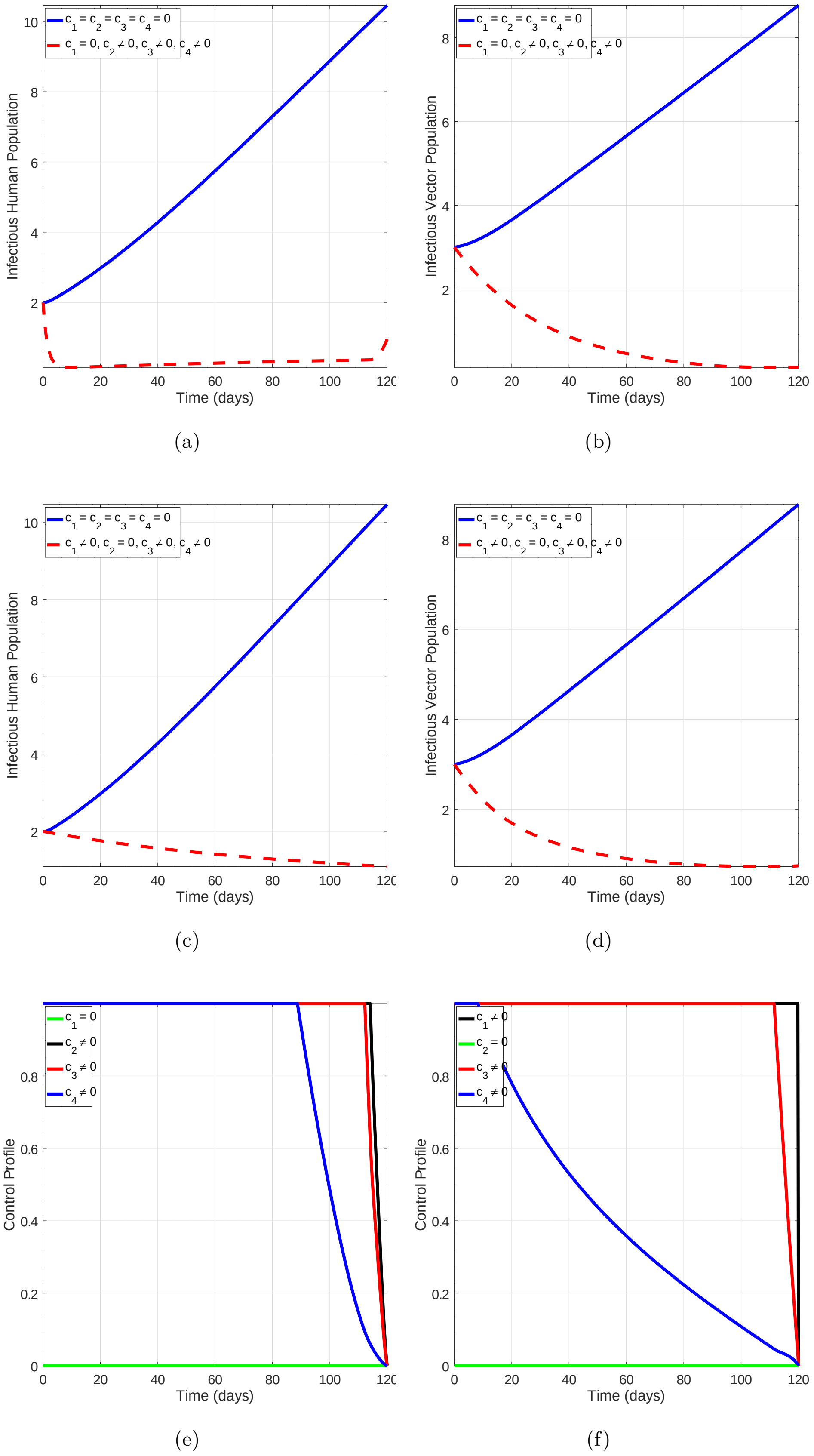


**Fig. A.6**: Simulations of the model (28) showing the efforts of intervention using three controls. (a) and (b) show the efforts of use of treatment, bed nets and insecticide spray on the infectious human population and infectious vector populations respectively. (c) and (d) show the efforts of using personal protection together, bed nets together with treatment on the infectious human population and infectious vector populations, respectively. Graphs (e)and (f) show control profiles for the corresponding control strategies.

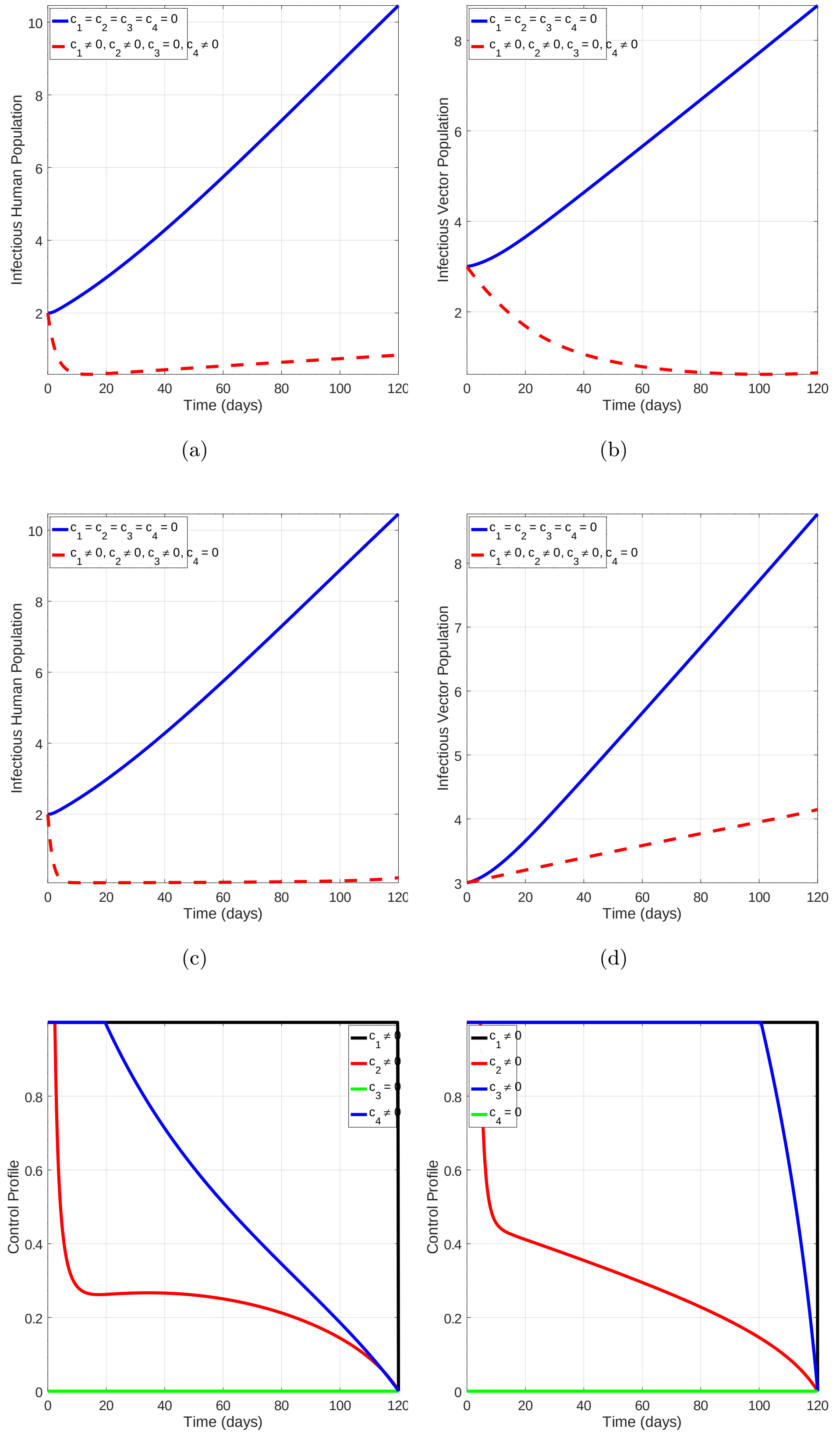


(a) (b)

(c) (d)

(e) (f)

**Fig. A.7**: Simulations of the model (28) showing the efforts of intervention using three controls. (a) and (b) show the combined efforts of treatment, use of personal protection gadgets and insecticide spray on the infectious human population and infectious vector populations respectively. (c) and (d) show the efforts of using personal protection, treatment and use of bed nets on the infectious human population and infectious vector populations, respectively. Graphs (e) and (f) show control profiles for the corresponding control strategies.

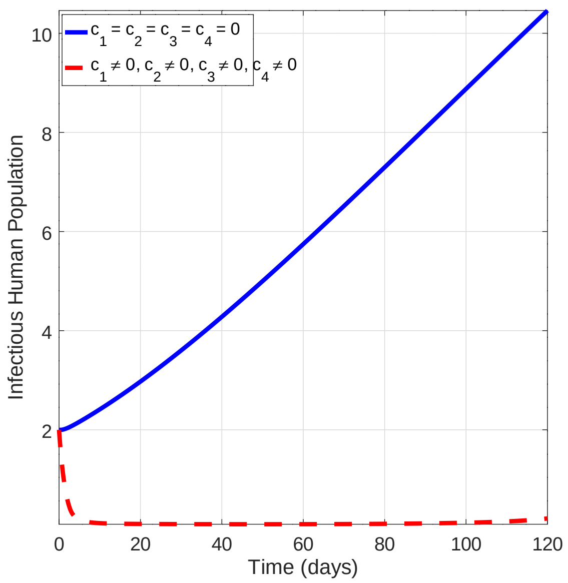


(a)

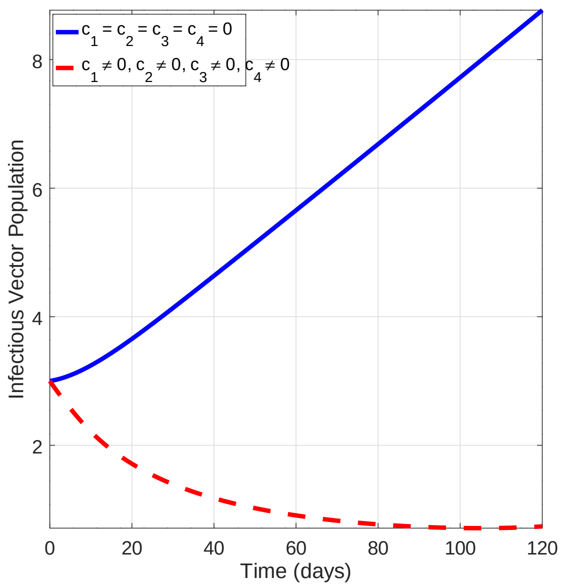


(b)

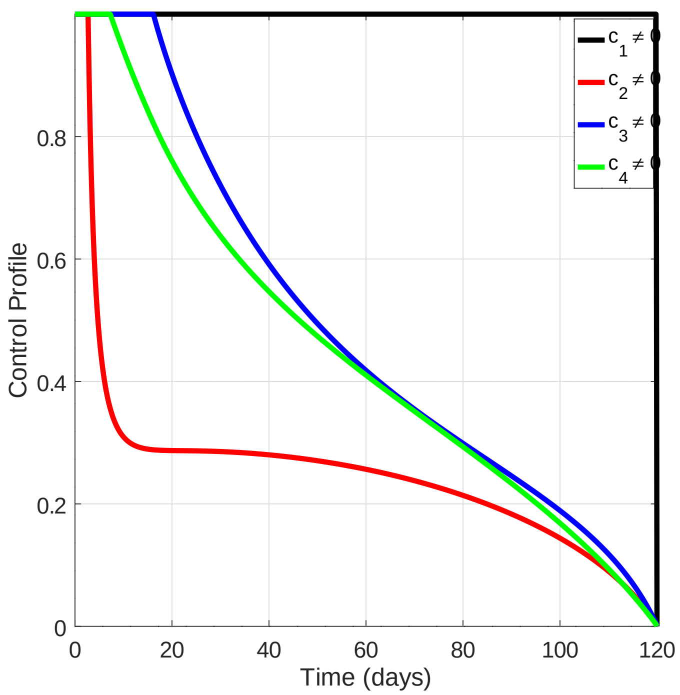


(c)

**Fig. A.8**: Model simulations showing the efforts of intervention by implementing all the four controls. (a) and (b) show the efforts of treatment, use of personal protection gadgets, bed nets and insecticide spray on the infectious human population and infectious vector populations, respectively. (c) shows control profiles for the corresponding control strategies.
